# The Women’s Wellness After Giving Birth Program (WWAGBP) for Vietnamese women: A single-arm trial protocol

**DOI:** 10.1101/2025.09.16.25335945

**Authors:** Lan Van Hoang, Thi Thanh Huong Nguyen, Ngoc Tran Tran, Thi Hoa Huyen Nguyen, Khanh Linh Bui, Debra Anderson, Kathleen Baird, Heena Akbar, Phuong Anh Hoang

## Abstract

**Background:** The transition to parenthood is a memorable yet vulnerable time for mothers’ physical and mental health, placing them at risk of postpartum depression. Prevalence of common perinatal mental disorders or postpartum depression among Vietnamese new mothers remains high, with potential long-term consequences for mothers’ mental health, children’s development, and family relationships. Supportive programs to prevent mental illness and promote mental health are, therefore, essential.

**Objective:** The 12-week women’s wellness after giving birth program (WWAGBP) aims to provide evidence-based knowledge and support for the mothers to reduce risk of postpartum depression while improving overall quality of life. This protocol outlines the design and planned methods of a pilot study to evaluate the feasibility and preliminary effectiveness of this health education package for postpartum Vietnamese women.

**Method:** Following ethical approval, 30 Vietnamese participants will be invited to a wellness program. Participants can access a free online platform featuring evidence-based information delivered through infographics, podcasts, pop-up key messages and videos. Primary outcomes include feasibility and usability (qualitative interviews and participant feedback). Secondary outcomes are risk of postpartum depression (Edinburgh Postnatal Depression Scale) and quality of life (SF-12), measured at 6 and 12 weeks.

**Results:** The WWAGBP protocol was finalized following funding approval in 2021, with program development completed within six months. Participant recruitment and pilot testing are projected to occur during the next six months, followed by preliminary data analysis for quantitative and qualitative findings.

**Conclusion:** This innovative intervention is the first of its kind in Vietnam to support women’s wellness after giving birth. Findings will help refine and optimize the program and provide valuable information on its effectiveness in promoting mental health, improving quality of life, and reducing the risk of postpartum depression among Vietnamese new mothers.

## INTRODUCTION

The transition to parenthood is a critical period for Vietnamese women after giving birth, during which they face a heightened vulnerability to common perinatal mental disorders and postpartum depression (PPD) (1, 2). PPD refers to depressive episodes within 4 weeks after birth, as specified by the Diagnostic and Statistical Manual of Mental Disorders, 5^th^ edition (3). However, it is essential to recognize that the trajectory of depressive symptoms can vary widely beyond this narrow time window (4, 5). Some individuals may experience milder “baby blues” symptoms that may progress to non-psychotic PPD while others may develop postpartum psychosis necessitating hospitalization (6, 7). PPD affects approximately 15% of new mothers globally, with variations observed across different regions (8-10). In Vietnam, the prevalence of PPD is even higher, standing at approximately 20% (11-13).

PPD is influenced by a range of risk factors, including personal and psychosocial factors such as a history of depression or anxiety, difficult childbirth experiences, lack of social support, stressful life events, marital discord, financial difficulties, premenstrual syndrome, and unintended pregnancy (10, 14-19). Hormonal fluctuations, particularly the rapid decline in reproductive hormones following birthing, have also been associated with the development of PPD (20-22). Postpartum depression and other perinatal mental disorders can have profound effects on the overall well-being of mothers. Studies exploring the lived experiences of new mothers have consistently highlighted symptoms such as persistent sadness, fatigue, feelings of being ignored, reduced interest in the baby, decreased appetite, and sleep disturbances among women experiencing PPD (23-26). A systematic review of qualitative evidence has revealed symptoms related to the inability to control feelings, ambivalence towards family members, imbalanced support between demands, and expression of hopelessness/helplessness (27). These conditions can lead to impaired maternal functioning, reduced maternal-infant bonding, and difficulties in daily caregiving tasks (28, 29). Untreated maternal mental health problems have been associated with adverse outcomes for children (30, 31), families (32) and even the intergenerational within families (33).

Primary care-based screening, diagnosis, and management have been effective in improving depression outcomes for mothers during the first 12 months (34). Non-psychotic PPD required proper care including timing screening, early interventions, and follow-up assessment in order not to worsen mental and physical health of women. The development and delivery of the 12-week wellness program are guided by the Theory of Planned Behavior (TPB), which posits that an individual’s behavior is directly influenced by their intention to perform that behavior, shaped by attitudes, subjective norms, and perceived behavioral control (35). In the context of maternal e-health, TPB-based interventions have effectively enhanced health behaviors by positively influencing these constructs. Web-based interventions, including mindfulness-based and peer support, have shown promise in preventing and addressing PPD (36-38). These interventions aim to enhance self-regulatory skills, such as emotion regulation, psychological flexibility, and self-compassion, which are crucial in the postpartum period (39).

In Vietnam, however, there has been insufficient maternal mental healthcare services for mothers during the postpartum period when women are at most risk of developing postpartum depression (34, 40). Overall, there has been a severe lack of maternal healthcare when it comes to providing care following birth and managing long-term postpartum depression. The continued use of unscientific traditional practices among women and family members following birth such as consulting fortune-tellers or following word-of-mouth remedies in certain areas of living has been identified as a challenge to evidence based postpartum care (41, 42). These practices can include avoiding bathing for 1 month as women’s physical health and body are believed to be too weak for bathing exposure, remaining in a dark room and avoiding bright lights in order not to suffer from sensitive and weak vision, using cotton swabs in ears to avoid tinnitus (43). Also, repeatedly eating foods that are said to stimulate lactation, such as pig’s feet, and avoid eating seafood as fishy taste of seafood can pass through breast milk and cause the baby to defecate. This poor diet, in contrast, turns out to affect mothers nutritional balance and lead to a loss of appetite following dietary restrictions, and by consuming special foods (43).

To address the unique challenges faced by Vietnamese women, it is crucial to understand the vulnerability of their mental health during the transition to parenthood and develop effective interventions to support their well-being. Interventions to improve maternal literacy during pregnancy, consultations scheduled for women after giving birth, and writing materials for references have been sporadically implemented in selected health care settings, however, mainly require physical attendance or scheduled arrangement. The prevalence of PPD among Vietnamese women has been noted as an increasing trend, which places it under high demands for further comprehensive and supportive programs targeting at postpartum women, their family members and the whole society. There should be a multi-approach program providing target audiences with knowledge and hands-on practices related to highly concerned topics after birth. The women should be benefited from evidence-based knowledge, up-to-date guidelines and practices following professional advice to be capable of undertaking selfcare for mothers and infants, reducing risk of postpartum mental disorders and improving their overall quality of life after birth. The digital interventions developed and tested across various contexts to support women’s wellness after childbirth were studied in other countries in the world. Findings from a pilot on 14 women proved the mobile application in the United States of America, which was designed to support the treatment of postpartum depression through cognitive behavioral therapy and interpersonal therapy techniques, to be acceptable, highly usable, and practical for weekly use in managing postpartum depression (44). In another research in China, an 8-week mobile health application integrating mindfulness and social support theories aimed at reducing postpartum depression symptoms resulted in significant improvements in maternal self-efficacy and perceived social support, along with a reduction in depressive symptoms (45).In Vietnam, with the high internet penetration rate, digital health innovations have the potential to reach a significant portion of the population (46). Finding a suitable number of sessions of appropriate durations and utilizing available functionality, interactivity, multimedia, and communication modes can maximize the long-term impact of internet-based interventions in postpartum women (47). This paper aims to describe a protocol to develop an internet-based program culturally tailored to address current challenges faced by Vietnamese women after giving birth.

## METHOD

### Overall aims of the program

The program overall aim is to examine the effectiveness and feasibility of a self-paced online-based health education intervention for new mothers.

### Study design

This is a single-arm trial composing a 12-week intervention period. The protocol was developed in accordance with the SPIRIT 2013 Statement guidelines for clinical trial protocols (48). Figure 1 demonstrates an overview of the protocol to develop and pilot the Women Wellness After Giving Birth program.

**Figure 1.**
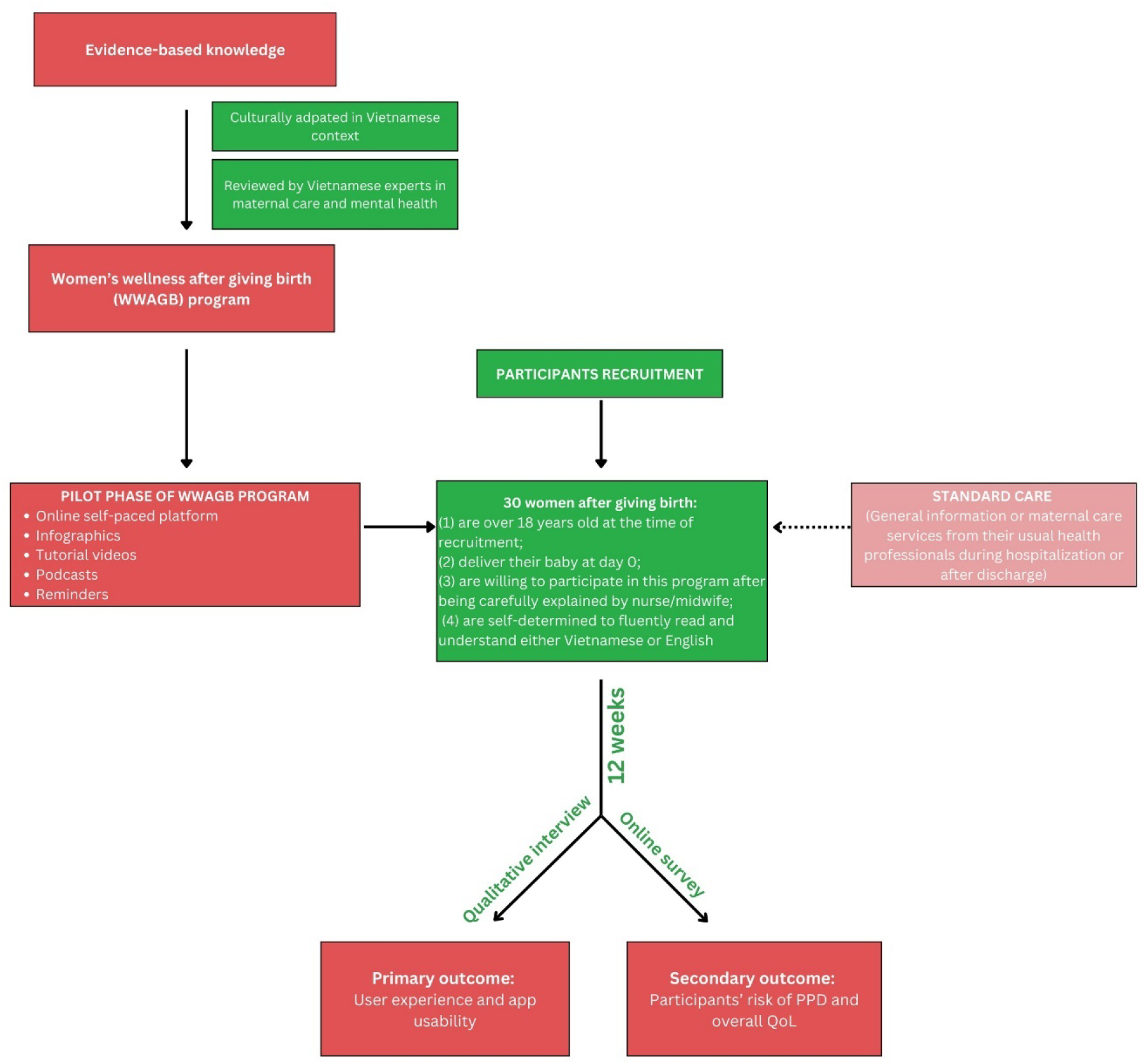
Overview of the protocol.

### The Women Wellness After Giving Birth (WWAGB) Program

WWAGB program is a 12-week self-paced web-based program targeting at new mothers to provide them with evidence-based information and interactive encouragement to both enhance their capacity to take care of themselves and their babies during the first 12 weeks and reduce risk of postpartum depression. Evidence-based knowledge refers to health information derived from peer-reviewed research, clinical guidelines, and expert consensus, culturally adapted for the Vietnamese postpartum population. Findings in the previous studies, for instance those applying similar interventions on postpartum women regarding the postpartum depression, share similar nuances that interventions with a duration of 12 weeks or more yielded more significant reductions in depressive symptoms compared to shorter programs (49, 50).

Additionally, the Interpersonal Psychotherapy, a validated treatment for postpartum depression, is typically structured over 12 to 16 weeks to allow for the establishment of a therapeutic alliance and the resolution of interpersonal issues contributing to depressive symptoms (51). While we acknowledge that the current program is not a treatment, the evidence suggests that a 12-week duration may be optimal for achieving meaningful clinical outcomes in postpartum mental health interventions. The idea to initiate content of 12-week program is based on the latest findings from researchers and health professionals worldwide focusing on promoting wellness and healthy behaviour change across women’s life, including physical activity, dietary intake, mental well-being; and to develop and test interventions that promote these behaviours. With that inspiration, the research team firstly conducted a survey to explore the current situation of mental wellness of Vietnamese women after birth and associated factors. Based on evidence from the exploratory study, the WWAGB program is then culturally developed in the Vietnamese context by the research team. Findings in the exploratory phase revealed common belief of Vietnamese women and their family members in caring for new mothers and the newborn, especially those living in the mountainous areas. Those unscientific traditional practices were used as references to develop content in relation to what a woman should and should not do. Additionally, in order to ensure the program is unique in the Vietnamese context, content of podcasts, infographics and videos were all proposed and co-contributed by the research team and health professionals who had years of experience working with Vietnamese women. The project then invited Vietnamese experts in maternal and mental health care including doctors, nurses, midwives, psychiatrists to review the related content before being finalized. A seri of workshops were conducted including content, technical and cultural adaptation to the Vietnamese context; technical delivery (website, eBook, printed handbook); and the utility and reliability of the WWAGBP package.

The educational package of WWAGB program has a range of informative and helpful topics for everyone either with or without a medical background including pregnant women, postpartum women, and women’s family members which has been written in a way and at a level that a non-medical background person could understand. The timing and application of these strategies were tailored to meet patient’s individual goals and functional capacities. Targeted health knowledge and behaviours cover 10 key topics including *Pain management; Postpartum physical changes; Hygiene; Baby feeding; Fatigue, Sleep and rest; Emotional wellbeing; Exercise; Diet and nutrition; Sexual life; and Social and emotional support*. Realizing that woman’s physical and psychological conditions are significantly changed after giving birth, and especially with the responsibility of caring for a newborn baby, the changes in the woman’s living habits are certain. The content of the program is designed and spread over first 12 weeks pospartum with the goal of becoming the woman’s companion and helping the woman to adapt and overcome difficulties in accordance with the specific changes of each week. Table 1 provides an example of how the package of content is structured.

**Table 1.** Example of WWAGB package of content and delivery method.

|  | Pain management | Postpartum physical changes | Hygiene | Baby feeding | Fatigue, sleep and rest | Emotional wellbeing | Exercise | Diet and nutrition | Sexual life | Social support and emotional support |
| --- | --- | --- | --- | --- | --- | --- | --- | --- | --- | --- |
| **Week 1** |  |  |  |  |  |  |  |  |  |  |
| Infographic | 1 | 1 | 1 | 1 |  | 1 |  |  |  |  |
| Podcast | 1 |  |  |  |  |  | 1 |  |  |  |
| Video |  |  |  |  |  |  | 1 |  |  |  |
| Pop-up key messages | 2 |  |  | 2 | 1 |  |  | 1 |  | 2 |
| **Week 8** |  |  |  |  |  |  |  |  |  |  |
| Infographic |  | 1 |  |  |  |  |  |  | 1 |  |
| Podcast |  |  |  |  |  |  |  |  |  |  |
| Video |  |  |  |  |  |  |  |  | 1 |  |
| Pop-up key messages |  |  |  | 1 |  | 1 | 2 |  |  | 1 |

*Note: Delivery formats (infographic, podcast, video, pop-up messages) were randomly assigned across weekly topics to optimize participant engagement and minimize content fatigue.*

The intervention content will be delivered through an interactive web interface which is friendly and flexible for women to enable self-learning at home or any preferred setting at their own suitable timeframe. The website will incorporate health-related news for women’s health; updated research evidence; an online community allowing participants to communicate with each other; and an overview of the program. Content of the 12-week program will be delivered to participants in the form of infographics, expert’s podcasts, pop-up key messages, and video tutorials, which are structured and tailored week by week to support mother’s journey. To enhance engagement and optimize user experience, different delivery formats will be randomly assigned to different content topics, helping to maintain participants’ interest and prevent monotonous repetition. Incorporating diverse multimedia formats—such as infographics, expert podcasts, pop-up key messages, and video tutorials—into postpartum wellness programs has been shown to enhance user engagement, comprehension, and behavior change. Infographics, for instance, simplify complex health information into visually appealing formats, improving knowledge retention and influencing health behaviors (52). Expert-led podcasts offer accessible, narrative-driven content that fosters behavioral change and social interaction, proving effective in promoting well-being and autonomy y(53). Pop-up key messages serve as timely reminders, reinforcing learning and encouraging adherence to health recommendations. Video tutorials provide dynamic, visual demonstrations that enhance understanding and skill acquisition, particularly beneficial for conveying practical health practices (54). To optimize women’s engagement throughout this 12 week, an email with recommended content of each day will be sent to women’s personal email to remind them throughout 12 weeks of the program. At the end of each delivered content such as infographic, podcast, pop-up key message or video, participants will be asked for their experience and feedback to improve the program in the future.

Rather than relying solely on expert opinion, the program content will be evaluated through direct feedback and health outcomes from new mothers who represent the intended users. The upcoming pilot phase as detailed in the following sections is proposed to validate content of the WWAGBP. After piloting with target audiences, content and delivery formats of the program will be re-assessed and adjusted upon the analysis of the research data.

### Study setting

The study is a self-paced online-based program piloted in Vietnam. Following the program activities, study settings will be adjustable, which includes but not limited to:

#### Virtual platform

The core designed activities will be delivered virtually throughout the program via the website where participants can flexibly self-learn in accordance with their schedule. Online self-determined assessment tools will be conducted at Week 6 and Week 12 for milestones screening of PDD and quality of life. This website can be easily launched on either smartphone or tablet or laptop.

#### Phone-based call

Participants will be followed up by a member of the research team via phone call to ask for user’s experience, feedback, and suggestions for future updates during pilot phase.

### Participant

The protocol to implement a finalized program will involve a trial phase before officially launching the program. As this is a pilot exploratory study, a sample size of 30 participants at Vinmec Times City International Hospital in Hanoi, Vietnam will be invited to participate to allow for initial assessment of the program’s feasibility, usability, and preliminary effectiveness. This size is considered adequate for identifying potential issues in intervention delivery and data collection procedures before scaling up to a larger trial. Women who (1) are over 18 years old at the time of recruitment, (2) deliver their baby at day 0, (3) are willing to participate in this program after being carefully explained by a nurse/midwife, and (4) are self-determined to fluently read and understand Vietnamese will be involved. Women will be excluded if they were currently diagnosed with severe postpartum complications preventing them from daily tasks, or had any clinical, cognitive or psychiatric contraindication identified by specialists in their medical records. These criteria will ensure participants completing the program to be able to follow suggested information without compromising their current health status.

### Recruitment process

The recruitment process will be multimodal to promote the program to potential participants. The offline promotion will be conducted at the Vinmec Times City International hospital. The research team will meet with key nurses and midwives at the hospital to introduce the program, targeted population, and all related benefits and drawbacks. These key staff will become research assistant to promote program to all women who are going to deliver at the hospital. Additionally, a presentation about research-related information will be introduced at a few antenatal classes for pregnant women and their family members who will become the potential participants in the program. All women waiting for their delivery or being at day 0 after giving birth at hospital will be explained about the program and invited to participate by a member of the research team. Using the information provided on the advertising material, participants can register for the program using a form created specifically for that purpose. This will enable participants to review the study material before completing the consent forms online.

### Standard care

At the same time participating in our program, as a client of the Vinmec Times City International hospital, participants in our project will keep being received standared maternal care services from their usual health professionals including from Department of Obstetrics and Gynaecology during hospitalization and from Department of Outpatients after discharge.

### Data collection

Data will be collected aligned with the outcomes of this research.

The primary outcome is to assess user experience, and app feasibility and usability, providing valuable insights for further program improvement.

- Methods: A qualitative approach will be applied to collect data for this primary outcome. Data will be initially collected from participant’s feedback after each content delivered on the web-based structured program. Additionally, researchers will conduct either an offline interview during scheduled postpartum visits or a phone call to participants after 12 weeks. A semi-structured recorded interview will be conducted in 30 to 45 minutes upon informed consent gained from participants.
- Data collection tool: Interview guide will be developed to provide participants with an open-ended question and prompts to structure the interview. The interview will start with a question asking *“What is your overall experience throughout 12 weeks of WWAGBP?”*. Prompts will be given in relation to content of WWAGBP, website interface, applicability, usability and feasibility. Data will be analyzed from participants’ feedback and recommendations regarding the course structure, content in each single activity, delivery strategies, communication methods and technical issues during their utilization.

The secondary outcome of this research is to evaluate the effectiveness of the program on maternal mental health and overall quality of life.

- Methods: Online survey integrated in the program will be used to assess participants’ risk of postpartum depression and overall quality of life after giving birth.
- Data collection tool: While the demographics information will be collected at the beginning of registration process, the self-assessment tool using Edinburgh Postnatal Depression Scale (EPDS) and the 12-Item Short-Form Health Survey (SF-12) has been attached to the program using a unique link for participants to click and response on their own preferred time at Week 6 and Week 12. The EPDS, with sensitivity and specificity of EPDS at 65–100% and 49–100% respectively, includes 10 items measured on a Likert scale of 0–3 (55). The validated Vietnamese version of EPDS is the most common screening tool for perinatal common mental disorders used in Vietnam with an internal consistency of 0.77 (56). Overall perceived quality of life will be assessed by Vietnamese-translated version of SF-12 (57), in which participants will be asked about the impact of physical and mental health on their everyday life during the last four weeks through 12 general health questions. Data will be summarized and extracted to SPSS 22.0 for analysis purposes by the research team after the pilot phase.

### Data analysis

For qualitative data, grounded theory methodology will be employed to systematically analyze participants’ narratives and identify key constructed concepts related to their experiences with the app. This approach allows for the generation of theory grounded in the data by engaging in iterative coding processes, including open, axial, and selective coding. The analysis will focus on uncovering patterns and themes that reflect users’ perceptions of the app’s usability, feasibility, and overall experience. Constant comparative analysis will be used to refine categories and explore relationships between them, ensuring that the findings remain closely tied to the participants’ lived experiences and the contextual factors influencing their engagement with the digital intervention.

For quantitative data, SPSS version 28.0 will be used to analyze the cleaned dataset. Descriptive statistics will first be conducted to summarize participant demographics and baseline characteristics. The key outcome variables will include the risk of PPD and quality of life among Vietnamese women who participate in the pilot phase of the program. Inferential statistical techniques, including regression analyses, will be applied to examine potential predictors and associations with the dependent variables. To strengthen the validity of the findings, potential confounding variables such as age, education level, employment status, parity, and pre-existing mental health conditions will be identified and recorded. Stratification will be employed during analysis to examine differences across key subgroups, while multivariable regression models will be used to control for these confounders when assessing the association between the intervention and changes in EPDS and SF-12 scores. Correlational analysis will also be used to explore the strength and direction of relationships between user engagement with the app and changes in health outcomes among vulnerable groups of participants. These analyses will help assess the effectiveness and potential impact of the digital intervention on participants’ well-being.

### Ethical consideration

- Human subject ethics review approvals or exemptions: This study received ethical approval from the Institutional Ethical Review Board of Vinmec International General Hospital JSC - VinUniversity (No 174/2021/QDD-VMEC dated 19^th^ October 2021).
- Informed consent: All participants will provide informed consent prior to participating in the study. Participants will be clearly explained why the pilot phase of this program is conducted and what it will involve as well as the risk and benefit to participate in the research. Before entering the online program, there will be a pop-up question to ensure the agreement of participant to participate. The willing to participate in the study is voluntary without any coercion and thus will not impact on any relationship between the participant and the research setting or between the participant and the research team member. All consent records will be securely stored on a password-protected server, accessible only to authorized research team members, in compliance with ethical standards for digital health research.
- Privacy and confidentiality: Data in the pilot phase of the current research will be anonymized and only accessed by the research team for research purpose. All the information will be reported as group information so that none of the personal data will be public. Data will be secured and stored in accordance with the university guidelines and policy.
- Compensation details: Participants will receive the reimbursement after they finish the questionnaire during the pilot process.

## RESULT

As this paper served to be a protocol for future projects to initiate research activities and propose timeline, thus key findings and outcomes from program activities will be disseminated in other publications instead. Following the program timeline, the Women’s Wellness After Giving Birth Program (WWAGBP) was funded under the VinUniversity Seed Funding scheme. By the submission date (September 2024), the project has successfully closed. Program development activities, including content creation, expert review, and technical platform building completed within six months.

Participant recruitment and data collection for pilot phase were followed by the end of 2022, corresponding to the 12-week intervention period and immediate follow-up. A total of 30 postpartum women were recruited to participate in the pilot study.

Preliminary data analysis has been initiated following the completion of data collection. Data analysis completed in 2023 and study results are expected to be disseminated in upcoming papers.

## DISCUSSION

The current paper explains the process of developing an internet-based program culturally tailored to address current challenges faced by Vietnamese new mothers. Acknowledging the necessity for healthcare professionals to pay more attention to prevent the risks of PPD and reduce consequences resulting from PPD, this paper provided an example of an comprehensive and smart approach to health education and self-care in maternal care. The WWAGB program is expected to be an evidence-based trustworthy resource contributing to delve into the intricacies of PPD situation in Vietnam.

To the best of our knowledge, the current paper presented a protocol for the first self-paced web-based program initiated for women after giving birth in Vietnam. In the era of information technology development and especially with the explosion of artificial intelligence, the online-based health education program seems to outweigh the original face-to-face health education delivery strategy, which equips target users with more convenient and timely access to resources, educational materials, and remote communication between healthcare providers and mothers (47).The intervention’s digital format inherently supports scalability, allowing the program to be expanded to a larger population of postpartum women without significant increases in resource demands. Similar to previous protocols for the development of health education programs which involved pilot phase as a key stage (58, 59), by piloting this WWAGB program, the remaining disadvantages of the internet-based delivery methods will be detected and revised for future implementations. Lessons learned from the COVID-19 pandemic necessitated the importance of the well-prepared resources for health promotion (60). In a similar nuance the expected program to be developed from the current protocol will mark a milestone for long-term development of e-health package and e-approach of health education, firstly in maternal care and then expand to other fields, which may remain effective during social distancing of other upcoming pandemics.. More significantly, the WWAGB program is promising to benefit a more diverse population including vulnerable groups in rural, remote and inaccessible places, as evidenced in a previous qualitative study indicating the advantages of using video-based approach to deliver maternal health education in mountainous areas in Vietnam(61).

In the current protocol to develop WWAGBP, the research team aimed to design a tailored health education package with infographics, podcasts, pop-up key messages and tutorial videos, which is expected to best fit postpartum women due to following reasons. Firstly, with a co-design approach, the expected content of the program has been validated and assured by different stages of review by national and international experts in maternal care and mental health care prior to the pilot phase. In order to ensure that educational interventions effectively deliver evidence-based knowledge to targeted participants, the content validity can help verify that the components of the intervention accurately reflect the intended knowledge. Interventions lacking content validity risk spreading knowledge that was not intended to be delivered, so undermining the potential positive impact of the intervention on desired participant’s outcomes (62). The co-design approach with a wide range of methods such as systematic review, qualitative research findings and expert knowledge as proposed in the current protocol has been previously proved its pivotal roles and effectiveness in the development of a web-based lifestyle intervention (63, 64). The information drawn from expertise of maternal and mental health and reviewed by national and international experts, therefore, is evidence and scientific-based for hands-on practices. Secondly, the flexibility of the program will help mothers self-schedule their time to be involved, which is remarkably different from postnatal consultation classes. Previous studies voiced the busy schedule and time constraint faced by women after giving birth due to their new babies and their tiredness (2, 23, 27, 29). Also, a web-based educational intervention without compulsory deadlines to finish learning activities will help ease the pressure on participants and prove their willingness to participate in their own preferred time. In a systematic review on women’s expectations and experiences regarding e-health treatment, the majority of women in previous studies has shown to be enticed by the convenience of the schedules offered by e-health in comparison to in-person program, which they found easier to integrate e-health interventions into their everyday life (65). Thirdly, new mothers always have to deal with a list of daily tasks preventing them from focusing on absorbing much information. As the overarching aim of e-Health interventions in postnatal education is not to fully replace face-to-face follow-up interactions, since they are crucial for health evaluations of the mother and infant by qualified health care providers (66). Also, findings from previous studies such as from a randomised control trial in South Korea regarding a postpartum depression self-care mobile app revealed that while the app was effective in improving mood, there were no significant differences in postpartum depression scores between the intervention and control groups (67). Nevertheless, this web-based interventions in the WWAGBP with an interactive format of information and condensed number of key messages can be utilized to supplement current postnatal care interactions in order to address areas of insufficient knowledge by delivering timely, standardised, pertinent, and evidence-based information directly to mothers.

By launching this WWAGB program, the research team expects to set up a protocol of developing e-health education package for women’s health across their life stages yet the current protocol may remain few limitations. As the WWAGBP will be the first internet-based program which is integrated different types of activities, the current paper may not anticipate all possible difficulties and challenges in the pilot phase. The proposed activities and content remains open to modify to best fit with target audiences yet commits to be assured by the professionals. The design which will not use two-arm randomized control trials may be controversial in the current protocol. However, with the aim to first time develop a novel approach to care for women after giving birth with respect to certain impacts of the current standard care, the research team consider the WWAGB as an additional evidence-based and interactive resource for new mothers in Vietnam. The effectiveness and feasibility of this self-paced web-based program will support further research projects targeting at raising awareness of the community and improving overall quality of life of women.

## CONCLUSION

Outcomes from this research project have the potential to impact on future clinical practices and e-health service delivery, especially on maternal care. WWAGB is a self-directed program aligning with and supporting the National Guidelines on Maternal care and standard postnatal care advice at the healthcare settings across Vietnam, which ends up improving quality of life for Vietnamese women after giving birth. This protocol will become the first officially reference for future development of similar and advance programs.

## ACKNOWLEDGMENT

We would like to express our deepest gratitude to the participants who generously shared their time and experiences with us. Our heartfelt thanks go to the research team at Vinmec Times City International Hospital, Hanoi, for their support and collaboration throughout this pilot study. Special thanks to the nursing and midwifery staff for their assistance in recruiting participants and promoting the program.

We are immensely grateful to the national and international experts in maternal care and mental health who provided critical insights and feedback during the development of the Women’s Wellness After Giving Birth Program.

We would also like to acknowledge the technical team responsible for developing the online platform.

Finally, we extend our sincere thanks to VinUniversity and the Institutional Ethical Review Board for their guidance and ethical oversight, ensuring that this study was conducted with the highest standards of research integrity.

## DATA AVAILABILITY

No datasets were generated or analyzed during the current study. Data resulting from the planned pilot trial will be made available upon reasonable request to the corresponding author after study completion, in accordance with PLOS ONE’s data sharing policy.

## AUTHOR CONTRIBUTIONS

LVH contributed to conceptualization, methodology, and writing – review and editing. TTHN contributed to conceptualization and methodology. NTT, THHN, and KLB contributed to project administration and writing – original draft. DA, KB, and HA provided supervision and contributed to writing – review and editing. PAH contributed to conceptualization and writing – review and editing.

## ABBREVIATIONS

WWAGBP: women’s wellness after giving birth program.

